# Tobacco control policies and smoking cessation treatment utilization: a moderated mediation analysis

**DOI:** 10.1101/2020.10.16.20213900

**Authors:** Johannes Thrul, Kira E. Riehm, Joanna E. Cohen, G. Caleb Alexander, Jon S. Vernick, Ramin Mojtabai

## Abstract

**Background:** Tobacco policies, including clean indoor air laws and cigarette taxes, increase smoking cessation in part by stimulating the use of cessation treatments. We explored whether the mediating effect of such treatments varies across socio-demographic groups.

**Methods:** We used data from 62,165 U.S. adult participants in the 2003 and 2010/11 Current Population Survey-Tobacco Use Supplement (CPS-TUS) who reported smoking cigarettes during the past year. Building on prior structural equation models used to quantify the degree to which smoking cessation treatments (prescription medications, nicotine replacement therapy, counselling/support groups, quitlines, and internet resources) mediated the association between clean indoor air laws, cigarette excise taxes, and recent smoking cessation, we added selected moderators to each model to investigate whether mediation effects varied by sex, race/ethnicity, education, income, and health insurance status.

**Results:** For clean indoor air laws, the mediating effect of prescription medication and nicotine replacement therapies varied significantly between racial/ethnic, age, and education groups in 2003. However, none of these moderation effects remained significant in 2010/11. For cigarette excise taxes in 2010/2011, the mediating effect of counseling was stronger in older adults; whereas, the mediating effect of prescription medications tended to be stronger in younger adults. No other moderator reached statistical significance. Smoking cessation treatments did not mediate the effect of taxes on smoking cessation in 2003 and were not included in these analyses.

**Conclusions:** Sociodemographic differences in how smoking cessation treatment use mediates between clean indoor air laws and smoking cessation have decreased from 2003 to 2010/11. In most cases, policies appear to stimulate smoking cessation treatment use similarly across varied sociodemographic groups.

## Introduction

Cigarette smoking is the single largest health risk behavior contributing to morbidity and mortality in the U.S. and responsible for more than 480,000 deaths each year (1). While quit attempts have increased and smoking prevalence has generally decreased over recent decades, there are entrenched differences in smoking prevalence among population subgroups (2). Members of racial/ethnic minority groups (2,3) and adults of low socioeconomic status (SES) (2,4) smoke cigarettes at higher rates than their counterparts. For example, 21.3% of adults with an annual household income <$35k smoke, compared to only 7.3% of those with an income of ≥$100k (2). Moreover, research has consistently shown differences in smoking prevalence by insurance status (5–7). Current smoking prevalence among Medicaid beneficiaries (23.9%) is more than twice that of privately insured individuals (10.5%) (2). As a consequence, smoking contributes to substantial health inequities, including marked disparities in cancer incidence, mortality, and cardiovascular disease risk in vulnerable populations (8,9). Better understanding of how to reduce smoking disparities is an urgent public health priority (10).

Tobacco control policies, including cigarette excise taxes and clean indoor air laws, have helped to reduce rates of cigarette smoking in the US (11–13). However, not all population subgroups may benefit equally from these policies. For example, while smokers of low socioeconomic status (SES) are just as likely to try quitting as other smokers, their attempts are less successful (1,14). Many low-SES smokers report past negative experiences with quitting (15) and have internalized smoking stigma, which may be associated with reduced self-efficacy for quitting (16). Additional unique obstacles to quitting faced by low-SES smokers include stronger nicotine dependence (4), social networks comprised of smokers, and strong pro-smoking social norms (17,18). With regards to racial/ethnic differences, adults who identified as African American, Hispanic, or other race/ethnicity report less use of pharmacotherapy for quitting smoking (19) and African American adults are less likely to attain successful short- or long-term smoking cessation compared to Whites (20).

We previously used the Current Population Survey-Tobacco Use Supplement (CPS-TUS) to demonstrate that U.S. smoking cessation increased from 2003 to 2010/11 and that changes in cigarette taxes and clean indoor air laws accounted for a substantial amount of this increase (21). Moreover, we found that cessation treatment use partly mediated the association of clean indoor air laws and smoking cessation in 2003 and the association of cigarette excise taxes and smoking cessation in 2010/11 (22). In the current study, we extend this work by examining whether the mediating effect of cessation treatments is similar across men and women, different age groups, racial/ethnic groups, and income levels, as well as groups with different health insurance coverage. Given that reducing smoking prevalence among vulnerable subpopulations is an urgent public health priority, this is a question of foremost importance for tobacco control research.

## Methods

### Sample

The CPS-TUS is a national population-level study of tobacco use conducted at regular intervals in conjunction with CPS. We used data from the 2003 and the 2010 and 2011 waves of the CPS-TUS. For 2003, the supplement was administered in February, June, and November 2003; for 2010 and 2011, TUS was administered in May and August 2010, and in January 2011. CPS uses a multi-stage stratified sampling procedure to interview a nationally representative sample of the non-institutionalized civilian U.S. population aged 15 years and older in 2003 and 18 years and older in 2010 and 2011. Approximately 64% of respondents complete the CPS-TUS by telephone and 36% in person. Most interviewees reported on their own tobacco use behavior; 20% reported as proxies for other household members. Additional information regarding the CPS-TUS can be found by visiting the TUS-CPS website (https://cancercontrol.cancer.gov/brp/tcrb/tus-cps/).

We limited our sample to past-year adult smokers, aged 18 and older, who reported on their own smoking behavior in CPS-TUS. A total of 34,842 participants in 2003 and 27,323 in 2010 and 2011 (for simplicity referred to as the 2011 CPS-TUS henceforth) met these criteria and were included.

### Measures

*Past year smoker* status was ascertained by asking participants about their smoking pattern exactly 12 months before the interview. This question was asked separately from current every-day and someday smokers as well as those who had quit in the past year. Individuals who reported smoking every day or on some days one year ago were rated as past-year smokers for this study.

*Quitting in the past year* was ascertained by responses “not at all” to the question “Do you now smoke cigarettes every day, some days, or not at all?” among past-year smokers. Among people who responded “not at all”, 82.7% had last smoked 30 days or longer before the time of interview.

*Cigarette excise taxes* and state and local *clean indoor air laws* were ascertained for each participant at the time point exactly one year before the time of their CPS-TUS interview. This timeframe was chosen because questions about smoking behavior in the CPS-TUS covered the past year. We obtained data on state and local cigarette excise taxes and clean indoor air laws from the American Nonsmokers’ Rights Foundation (ANRF). Total excise tax was computed as the sum of federal, state, and local taxes. ANRF ascertains data on state and local clean indoor air laws separately for laws affecting workplace areas, bars, and restaurants. We used the ANRF categorization of these laws into those imposing a “100% smoke free policy,” a “qualified 100% smoke free policy,” laws providing “some” coverage, and “no coverage”. In situations where the state and local laws affecting a participant were inconsistent, we chose the more comprehensive law. While some states pre-empt, or disallow, local tobacco control laws, the number of such pre-emptive state laws affecting clean indoor air policies decreased over the study period: 12 states had such laws in 2010, down from 18 in 2000 (23). State and local law data were linked to the CPS-TUS data using state and county FIPS codes. Because state and local laws affecting workplace, bars, and restaurants are strongly correlated (r range=.61 to .83), an average clean indoor law index was computed.

*Smoking cessation treatment use* was assessed by asking past-year smokers who had quit or had made an attempt to quit about the methods they had used. These methods included nicotine replacement treatments (nicotine gum, lozenge, patch, inhaler, or nasal spray), prescription pills (Zyban, Wellbutrin, or bupropion; Chantix or varenicline was added in 2010/11), telephone help line or quit line, one-on-one counseling, stop smoking clinic, class, or support group (combined into a “counseling/groups”), and the internet.

*Family income* and *health insurance coverage* were assessed in the 2003, 2010 and 2011 Annual Social and Economic (ASEC) Supplement of CPS that is administered in March of each year and has partial overlap with the CPS-TUS sample (24). As such, for a smaller proportion of CPS-TUS participants information on income and health insurance is available. For this study, annual family income was categorized into three categories (<20,000$, 20,000$ - <75,000$, and ≥75,000$) and health insurance into 3 mutually exclusive groups: public insurance (Medicare, Medicaid, VA/CHAMPS, Indian Health Services), private insurance (either through job or personally obtained) with or without public insurance; and no health insurance.

In addition to questions about smoking and smoking cessation treatments, CPS-TUS also collected socio-demographic data including sex, age (18-29, 30-49, 50-64, 65+), race/ethnicity (non-Hispanic white, non-Hispanic black, Hispanic, other), education (< High School, High School graduate or GED, some college but not bachelor’s degree, bachelor’s degree or higher), employment status (employed, unemployed, not in labor force), and marital status (married or living as married, widowed, divorced or separated, never married). We also adjusted the analyses for country region and state-level expenditure for tobacco prevention programs compared to US Centers for Disease Control and Prevention (CDC) recommended expenditure for years 2003 and 2011 compiled by the Campaign for Tobacco-Free Kids (https://www.tobaccofreekids.org/assets/factsheets/0209.pdf).

### Analyses

We analyzed the data in two stages. First, we examined variations in the use of different smoking cessation treatments across socio-demographic and health insurance groups using contingency tables.

Second, we conducted moderated mediation analysis (Figure 1) using structural equation modeling with binary outcomes and multiple binary mediators (25,26) to examine the extent to which mediated effects between clean indoor air laws and taxes, on the one hand, and recent quitting, on the other hand, varied across socio-demographic groups. Because both the mediators (different treatments) and most of the moderating factors were categorical, we conducted these moderated mediation analyses by multi-group structural equation modeling. All coefficients in these models were fixed to be constant across groups except for the coefficients linking the exposure (clean indoor air laws in 2003; clean indoor air laws or taxes in 2010/11) to each mediator. These latter coefficients were allowed to vary across groups (27). Moderated effects were tested by comparing these coefficients for each treatment across groups. To avoid spurious findings, moderation was tested for preselected socio-demographic variables that were associated with smoking cessation treatment use in previous research, and these variables included sex, race/ethnicity, age, education, income and health insurance. These models also adjusted for employment status, marital status, country region, and state-level expenditure for tobacco prevention programs compared to CDC recommendations. All analyses were conducted separately for the 2003 period and the 2010/11 period. These analyses build on our previous set of analyses that examined whether and to what extent cessation treatment policies mediated the effect of clean indoor air laws and cigarette excise taxes on recent smoking cessation (22). Analyses for 2003 were limited to the effect of clean indoor air laws because only these laws were associated with smoking cessation in 2003. Analyses for 2010/11 included both clean indoor air laws and cigarette excise taxes, as cessation treatments mediated the effect of both (22).

**Figure 1.**
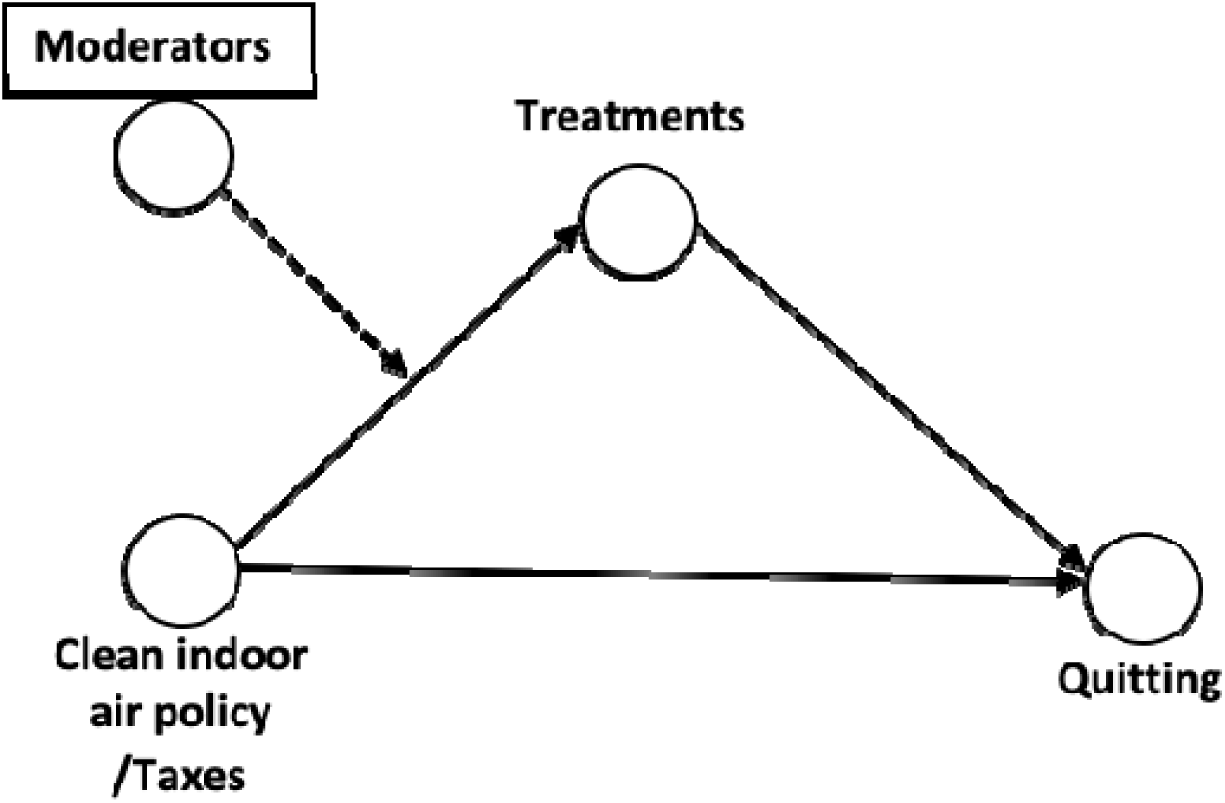

Analyses were conducted using Stata 16.0 software (StataCorp LLC, College Station, TX, 2019). Structural equation modeling analyses were conducted using the *gsem* routine of Stata which accommodates binary outcomes and mediators as well complex survey data with *successive difference replications* as required for analyses of census data (28). Survey and replicate weights were included in all analyses to compute population representative estimates and confidence intervals. All percentages reported are weighted. A conservative p<0.01 cutoff was used for deciding the statistical significance of the tests.

## Results

### Sample characteristics

We previously described characteristics of past-year smokers in the 2003 and the 2010/11 samples (21,22). Briefly, the majority of participants in both time periods were male (53.7% in 2003 and 54.1% in 2010/11), non-Hispanic white (75.7% and 74.4%), and employed (66.0% and 58.6%). The average age of the participants was 41.41 years (standard error [SE]=.05) in 2003 and 42.69 years (SE=0.10) in 2010/11. The proportion married or living as married were 43.7% in 2003 and 39.9% in 2010/11. The South region had the largest proportion of participants in both set of samples (37.5% in 2003 and 39.3% in 2010/11), and the Northeast region had the smallest proportion (18.0% and 16.4%, respectively).

### Tobacco control policies

We have also previously reported variations in clean indoor air laws and taxes in the two time periods (21,22). State and local governments varied considerably in their adoption of tobacco control policies and the extent of coverage changed markedly over time. In 2003, only 1.9% of past-year smokers lived in states and localities with 100% smoke-free workplace laws, 8.2% in states and localities with 100% smoke-free bar clean indoor air laws and 9.0% in states and localities with 100% smoke-free restaurant laws. These numbers increased to 47.7%, 44.3% and 53.5%, respectively, in 2010/11. Excise taxes also increased over time from an average of $1.00 (SE=.001) to $2.25 (SE=.005). The proportion of past-year smokers who quit did not change much over two periods: 7.3% in 2003 and 7.8% in 2010/11 quitted smoking. The most commonly used treatments in both 2003 and 2010/11 were nicotine replacement therapies, used by 10.3% of past-year smokers in 2003 and 2010/11, followed by prescription medications, used by 3.9% in 2003 and 5.9% in 2010/11 (Tables 1 and 2).

**Table 1:** Use of smoking cessation treatments and smoking cessation in 34,842 participants Current Population Survey-Tobacco Use Supplement, 2003, according to sociodemographic characteristics and type of treatment.

| Groups | N | Prescription medications | Replacement therapies | Counselling/ Groups | Quitline | Internet |
| --- | --- | --- | --- | --- | --- | --- |
|  |  | N (%) | N (%) | N (%) | N (%) | N (%) |
| Total | 34842 | 1508 (3.9) | 3877 (10.3) | 518 (1.3) | 233 (0.5) | 217 (0.6) |
| Sex |  |  |  |  |  |  |
| Male | 16792 | 581 (3.1) | 1731 (9.5) | 197 (1.0) | 74 (0.4) | 78 (0.5) |
| Female | 18050 | 927 (4.8) | 2146 (11.2) | 321 (1.6) | 159 (0.7) | 139 (0.8) |
| Comparison | | $X^2_{df=1}=184.01$,<br>p<0.001 | $X^2_{df=1}=84.00$,<br>p<0.001 | $X^2_{df=1}=99.34$,<br>p<0.001 | $X^2_{df=1}=63.86$,<br>p<0.001 | $X^2_{df=1}=48.24$,<br>p<0.001 |
| Race/ethnicity |  |  |  |  |  |  |
| Non-Hispanic white | 27862 | 1339 (4.5) | 3311 (11.2) | 427 (1.4) | 186 (0.5) | 186 (0.7) |
| Non-Hispanic black | 2879 | 55 (1.8) | 220 (7.1) | 37 (1.1) | 20 (0.7) | 10 (0.3) |
| Hispanic | 2180 | 32 (1.2) | 158 (6.5) | 17 (0.6) | 11 (0.5) | 10 (0.5) |
| Other | 1921 | 82 (3.4) | 188 (9.2) | 37 (1.5) | 16 (0.6) | 11 (0.5) |
| Comparison | -- | $X^2_{df=3}=387.92$,<br>p<0.001 | $X^2_{df=3}=257.51$,<br>p<0.001 | $X^2_{df=3}=44.87$,<br>p<0.001 | $X^2_{df=3}=8.33$,<br>p=0.040 | $X^2_{df=3}=24.65$,<br>p<0.001 |
| Age, years |  |  |  |  |  |  |
| 18-29 | 7348 | 174 (2.0) | 576 (7.0) | 56 (0.6) | 56 (0.6) | 52 (0.7) |
| 30-49 | 16050 | 772 (4.3) | 1916 (11.1) | 229 (1.2) | 103 (0.4) | 111 (0.7) |
| 50-64 | 8172 | 469 (5.8) | 1089 (13.1) | 176 (1.9) | 53 (0.6) | 50 (0.7) |
| 65+ | 2913 | 89 (3.1) | 272 (9.1) | 43 (1.6) | 18 (0.7) | 4 (0.2) |
| Comparison | | $X^2_{df=3}=397.06$,<br>p<0.001 | $X^2_{df=3}=427.88$,<br>p<0.001 | $X^2_{df=3}=248.42$,<br>p<0.001 | $X^2_{df=3}=14.18$,<br>p=0.003 | $X^2_{df=3}=14.29$,<br>p=0.003 |
| Education |  |  |  |  |  |  |
| <HS gradate | 6403 | 181 (2.5) | 570 (7.8) | 69 (0.9) | 38 (0.5) | 11 (0.1) |
| HS graduate or GED | 13947 | 563 (3.6) | 1402 (9.3) | 168 (1.0) | 74 (0.4) | 48 (0.3) |
| College, < bachelor's degree | 10089 | 540 (5.0) | 1280 (12.0) | 193 (1.7) | 94 (0.8) | 98 (1.0) |
| Bachelor's degree or more | 4403 | 224 (4.8) | 625 (13.3) | 88 (1.7) | 27 (0.5) | 60 (1.4) |
| Comparison | | $X^2_{df=3}=208.29$,<br>p<0.001 | $X^2_{df=3}=389.00$,<br>p<0.001 | $X^2_{df=3}=91.47$,<br>p<0.001 | $X^2_{df=3}=52.83$,<br>p<0.001 | $X^2_{df=3}=267.57$,<br>p<0.001 |
| Employment |  |  |  |  |  |  |
| Employed | 22822 | 999 (3.9) | 2563 (10.3) | 290 (1.1) | 137 (0.4) | 158 (0.7) |
| Unemployed | 2480 | 83 (2.8) | 230 (8.6) | 38 (0.9) | 16 (0.5) | 15 (0.7) |
| Not in labor force | 9540 | 426 (4.2) | 1084 (10.6) | 190 (1.8) | 80 (0.7) | 44 (0.4) |
| Comparison | | $X^2_{df=2}=29.58$,<br>p<0.001 | $X^2_{df=2}=23.07$,<br>p<0.001 | $X^2_{df=2}=86.17$,<br>p<0.001 | $X^2_{df=2}=33.40$,<br>p<0.001 | $X^2_{df=2}=28.38$,<br>p<0.001 |
| Marital status |  |  |  |  |  |  |
| Married/living as married | 15854 | 818 (4.8) | 1943 (11.5) | 259 (1.4) | 107 (0.5) | 99 (0.6) |
| Widowed | 1806 | 72 (3.8) | 178 (9.1) | 24 (1.3) | 8 (0.4) | 6 (0.4) |
| Divorced/separated | 8015 | 385 (4.6) | 966 (11.9) | 123 (1.3) | 63 (0.6) | 53 (0.7) |
| Never married | 9167 | 233 (2.2) | 790 (7.6) | 112 (1.0) | 55 (0.5) | 59 (0.7) |
| Comparison | | $X^2_{df=3}=255.55,$<br>$p<0.001$ | $X^2_{df=3}=272.45,$<br>$p<0.001$ | $X^2_{df=3}=27.95,$<br>$p<0.001$ | $X^2_{df=3}=5.78,$<br>$p=0.123$ | $X^2_{df=3}=7.32,$<br>$p=0.062$ |
| Family income<sup>a</sup> |  |  |  |  |  |  |
| <\$20K | 4774 | 154 (2.7) | 435 (8.4) | 62 (1.0) | 38 (0.6) | 22 (0.5) |
| \$20-<\$75K | 7748 | 404 (4.6) | 944 (11.3) | 120 (1.3) | 53 (0.6) | 45 (0.5) |
| \$75K+ | 1896 | 114 (5.5) | 270 (13.7) | 30 (1.6) | 8 (0.3) | 24 (1.5) |
| Comparison | | $X^2_{df=2}=156.16,$<br>$p<0.001$ | $X^2_{df=2}=224.84,$<br>$p<0.001$ | $X^2_{df=2}=14.41,$<br>$p<0.001$ | $X^2_{df=2}=7.58,$<br>$p=0.023$ | $X^2_{df=2}=73.41,$<br>$p<0.001$ |
| Health insurance<sup>a</sup> |  |  |  |  |  |  |
| Private | 8205 | 459 (5.1) | 1052 (12.1) | 126 (1.4) | 55 (0.5) | 64 (0.8) |
| Public | 2563 | 104 (3.9) | 284 (11.1) | 57 (2.0) | 29 (1.1) | 13 (0.6) |
| None | 2870 | 77 (2.1) | 223 (6.5) | 25 (0.6) | 12 (0.2) | 8 (0.2) |
| Comparison | | $X^2_{df=2}=149.62,$<br>$p<0.001$ | $X^2_{df=2}=252.03,$<br>$p<0.001$ | $X^2_{df=2}=61.12,$<br>$p<0.001$ | $X^2_{df=2}=46.16,$<br>$p<0.001$ | $X^2_{df=2}=28.45,$<br>$p<0.001$ |

a. Family income and health insurance coverage were assessed in the Annual Social and Economic (ASEC) Supplement of CPS administered in March 2003 and has partial overlap with the CPS-TUS sample. As such, this information is not available for the full CPS-TUS sample.

**Table 2:** Use of smoking cessation treatments and smoking cessation in 27,323 participants Current Population Survey-Tobacco Use Supplement, 2010/11, according to sociodemographic characteristics and type of treatment.

| **Groups** | **N** | **Prescription**<br>**medications**<br>**N (%)** | **Replacement**<br>**therapies**<br>**N (%)** | **Counselling/**<br>**Groups**<br>**N (%)** | **Quitline**<br>**N (%)** | **Internet**<br>**N (%)** |
| --- | --- | --- | --- | --- | --- | --- |
| Total | 27323 | 1866 (5.9) | 3066 (10.3) | 555 (1.7) | 482 (1.3) | 236 (0.8) |
| Sex |  |  |  |  |  |  |
| Male | 13538 | 745 (4.6) | 1414 (9.4) | 215 (1.4) | 168 (1.0) | 88 (0.6) |
| Female | 13785 | 1121 (7.4) | 1652 (11.3) | 340 (2.0) | 314 (1.7) | 148 (1.1) |
| Comparison | -- | $X^2_{df=1}=123.11$,<br>$p<0.001$ | $X^2_{df=1}=31.56$,<br>$p<0.001$ | $X^2_{df=1}=16.48$,<br>$p<0.001$ | $X^2_{df=1}=44.38$,<br>$p<0.001$ | $X^2_{df=1}=19.04$,<br>$p<0.001$ |
| Race/ethnicity |  |  |  |  |  |  |
| Non-Hispanic white | 21263 | 1629 (6.8) | 2512 (10.9) | 419 (1.7) | 395 (1.4) | 200 (0.9) |
| Non-Hispanic black | 2587 | 97 (3.2) | 237 (9.0) | 76 (2.6) | 39 (1.1) | 10 (0.4) |
| Hispanic | 1857 | 50 (2.4) | 143 (6.8) | 22 (1.0) | 23 (1.1) | 11 (0.5) |
| Other | 1616 | 90 (5.1) | 174 (10.6) | 38 (1.4) | 25 (1.2) | 15 (1.2) |
| Comparison | -- | $X^2_{df=3}=118.96$,<br>$p<0.001$ | $X^2_{df=3}=44.79$,<br>$p<0.001$ | $X^2_{df=3}=27.50$,<br>$p<0.001$ | $X^2_{df=3}=2.59$,<br>$p=0.460$ | $X^2_{df=3}=10.37$,<br>$p=0.016$ |
| Age, years |  |  |  |  |  |  |
| 18-29 | 5306 | 157 (2.5) | 457 (7.6) | 74 (1.0) | 87 (1.0) | 64 (1.2) |
| 30-49 | 11241 | 827 (6.6) | 1315 (11.1) | 214 (1.7) | 201 (1.4) | 96 (0.7) |
| 50-64 | 8080 | 679 (7.6) | 1001 (11.6) | 217 (2.4) | 158 (1.7) | 63 (0.7) |
| 65+ | 2696 | 203 (7.1) | 293 (10.3) | 50 (1.5) | 36 (0.9) | 13 (0.5) |
| Comparison | | $X^2_{df=3}=143.90$,<br>$p<0.001$ | $X^2_{df=3}=68.37$,<br>$p<0.001$ | $X^2_{df=3}=66.09$,<br>$p<0.001$ | $X^2_{df=3}=21.13$,<br>$p<0.001$ | $X^2_{df=3}=18.51$,<br>$p<0.001$ |
| Education |  |  |  |  |  |  |
| <HS graduate | 4428 | 227 (4.2) | 423 (8.2) | 90 (1.5) | 67 (1.1) | 21 (0.4) |
| HS graduate or GED | 10920 | 698 (5.6) | 1120 (9.5) | 185 (1.4) | 163 (1.0) | 54 (0.5) |
| College, < bachelor's degree | 8538 | 710 (7.1) | 1077 (11.7) | 203 (2.0) | 192 (1.8) | 102 (1.3) |
| Bachelor's degree or more | 3437 | 231 (6.2) | 446 (12.2) | 77 (1.9) | 60 (1.5) | 59 (1.5) |
| Comparison | | $X^2_{df=3}=68.98$,<br>$p<0.001$ | $X^2_{df=3}=57.76$,<br>$p<0.001$ | $X^2_{df=3}=13.71$,<br>$p=0.003$ | $X^2_{df=3}=25.36$,<br>$p<0.001$ | $X^2_{df=3}=56.11$,<br>$p<0.001$ |
| Employment |  |  |  |  |  |  |
| Employed | 15906 | 1045 (5.6) | 1724 (10.1) | 268 (1.4) | 246 (1.1) | 142 (0.8) |
| Unemployed | 2908 | 162 (4.8) | 295 (8.6) | 49 (1.3) | 57 (1.4) | 30 (1.0) |
| Not in labor force | 8509 | 659 (6.8) | 1047 (11.4) | 238 (2.4) | 179 (1.6) | 64 (0.7) |
| Comparison | | $X^2_{df=2}=25.28$,<br>$p<0.001$ | $X^2_{df=2}=22.26$,<br>$p<0.001$ | $X^2_{df=2}=47.29$,<br>$p<0.001$ | $X^2_{df=2}=10.58$,<br>$p=0.005$ | $X^2_{df=2}=2.85$,<br>$p=0.240$ |
| Marital status |  |  |  |  |  |  |
| Married/living as married | 11380 | 954 (7.6) | 1303 (10.7) | 228 (1.7) | 181 (1.3) | 107 (0.9) |
| Widowed | 1527 | 123 (6.9) | 196 (12.2) | 39 (2.7) | 35 (2.0) | 8 (0.4) |
| Divorced/Separated | 6649 | 493 (6.6) | 809 (11.4) | 164 (2.1) | 148 (1.7) | 51 (0.7) |
| Never married | 7767 | 296 (3.1) | 758 (8.6) | 124 (1.2) | 118 (1.0) | 70 (0.9) |
| Comparison | | $X^2_{df=3}=214.87,$<br>$p<0.001$ | $X^2_{df=3}=54.04,$<br>$p<0.001$ | $X^2_{df=3}=32.19,$<br>$p<0.001$ | $X^2_{df=3}=17.14,$<br>$p<0.001$ | $X^2_{df=3}=4.11,$<br>$p=0.250$ |
| Family income<sup>a</sup> |  |  |  |  |  |  |
| <\$20K | 7717 | 438 (4.8) | 869 (9.8) | 194 (1.9) | 179 (1.7) | 48 (0.6) |
| \$20-<\$75K | 15216 | 1057 (5.9) | 1672 (10.3) | 294 (1.6) | 249 (1.2) | 130 (0.8) |
| \$75K+ | 4390 | 371 (7.8) | 525 (11.0) | 67 (1.4) | 54 (1.1) | 58 (1.3) |
| Comparison | | $X^2_{df=2}=62.70$<br>$p<0.001$ | $X^2_{df=2}=4.94,$<br>$p=0.085$ | $X^2_{df=2}=4.42,$<br>$p=0.110$ | $X^2_{df=2}=15.66,$<br>$p<0.001$ | $X^2_{df=2}=15.44,$<br>$p<0.001$ |
| Health insurance<sup>a</sup> |  |  |  |  |  |  |
| Private | 5786 | 473 (7.2) | 674 (11.1) | 115 (1.7) | 98 (1.4) | 56 (0.8) |
| Public | 2854 | 220 (6.7) | 361 (12.4) | 77 (2.6) | 70 (2.1) | 27 (1.0) |
| None | 2814 | 91 (2.7) | 251 (8.2) | 35 (0.8) | 34 (0.9) | 17 (0.6) |
| Comparison | | $X^2_{df=2}=68.98,$<br>$p<0.001$ | $X^2_{df=2}=31.92,$<br>$p<0.001$ | $X^2_{df=2}=36.86,$<br>$p<0.001$ | $X^2_{df=2}=16.00,$<br>$p<0.001$ | $X^2_{df=2}=3.12,$<br>$p=0.210$ |

a. Family income and health insurance coverage were assessed in the Annual Social and Economic (ASEC) Supplement of CPS administered in March 2003 and has partial overlap with the CPS-TUS sample. As such, this information is not available for the full CPS-TUS sample.

### Variations in the use of smoking cessation services across population groups

There were significant sociodemographic variations in the use of smoking cessation treatments. Women were consistently more likely to use all forms of treatment both in 2003 and 2010/11. Similarly, non-Hispanic whites were more likely to use most forms of treatment in both periods, with the few exceptions of use of quitlines in both 2003 and 2010/11, use of internet in 2010/11, as well as use of counseling and groups by “other” racial/ethnic groups in 2003 and non-Hispanic blacks in 2010/11.

There were some consistencies in the patterns of use of services according to age as well. Compared to other age groups, adults in the 50-64 years age range were most likely and those in the 18-29 years age group least likely to use prescription medications, replacement therapies and counseling and groups in both 2003 and 2010/11 periods.

Adults with higher education were more likely than those with less education in both periods to use all services, a gradient in use of services according to family income was also found for prescription medications and replacement therapies in both 2003 and 2010/11 and for counseling and groups in 2003: individuals with family income <$20,000 were least likely to use these services; whereas, those with family incomes ≥ $75,000 were most likely. Individuals with no health insurance coverage, compared to those with private and public insurance, were less likely to use any kind of treatment in both periods, including treatments that are typically free of charge or not reimbursed by health insurance, such as internet resources and quitlines. The associations with marital status were less consistent.

### Moderated mediation analyses

We limited our moderated mediation analyses to prescription medications and nicotine replacement therapies for mediation analyses of clean indoor air laws in 2003 and 2010/11 because only these treatments mediated the association of clean indoor air laws with recent smoking cessation in both periods (22). For the same reason, we restricted our moderated mediation analyses for taxes in 2010/11 to prescription medications, nicotine replacement therapies, and counseling/group therapy in 2010/11 (22). There were variations in mediated effects across socio-demographic groups in both 2003 and 2010/11, although some of the mediation coefficients did not reach a statistically significant level due to small number of participants in some demographic groups who used smoking cessation treatments (e.g., racial/ethnic minorities).

The mediating effects of prescription medications and nicotine replacement therapies for the association of clean indoor air laws with quitting were most prominent in non-Hispanic whites and blacks in 2003. Furthermore, the mediating effect of treatments were larger in the age groups 50-64 and 30-49 in 2003 (Table 3). However, such racial/ethnic and age variations were not apparent in 2010/11. The mediating effects of prescription medications for the association of clean indoor air laws and smoking cessation in 2003 were somewhat smaller among those who had not graduated high school and those with advanced degrees. A similar pattern was noted with regard to higher education and use of replacement therapies. However, the education effects were not apparent in 2010/11.

**Table 3:** Mediating effect of smoking cessation treatments in the association of clean indoor air policies and smoking cessation in different socio-demographic groups (moderated mediation), 2003 and 2010/11.^a^

| Group | 2003 survey |  | 2010/11 survey |  |
| --- | --- | --- | --- | --- |
|  | Prescription medications | Replacement Therapies | Prescription medications | Replacement therapies |
| Overall | 1.08 (1.02-1.13) | 1.11 (1.07-1.15) | 1.08 (1.03-1.14) | 1.09 (1.05-1.13) |
| Sex |  |  |  |  |
| Male | 1.09 (1.00-1.19) | 1.07 (1.02-1.13) | 1.06 (0.98-1.14) | 1.05 (0.99-1.12) |
| Female | 1.08 (1.00-1.16) | 1.04 (0.99-1.09) | 1.06 (1.00-1.13) | 1.01 (0.96-1.06) |
| Comparison of groups | $X^2_{df=1}=0.06$,<br>p=0.812 | $X^2_{df=1}=1.05$,<br>p=0.306 | $X^2_{df=1}=0.00$,<br>p=0.973 | $X^2_{df=1}=2.13$,<br>p=0.145 |
| Race/ethnicity |  |  |  |  |
| Non-Hispanic white | 1.14 (1.06-1.22) | 1.09 (1.04-1.14) | 1.06 (1.00-1.13) | 1.02 (0.97-1.06) |
| Non-Hispanic black | 1.12 (0.87-1.45) | 1.28 (1.16-1.42) | 1.06 (0.89-1.26) | 1.09 (0.96-1.25) |
| Hispanic | 0.61 (0.43-0.85) | 0.83 (0.72-0.95) | 0.98 (0.72-1.34) | 1.15 (0.98-1.35) |
| Other | 0.80 (0.69-0.92) | 0.83 (0.73-0.94) | 1.06 (0.85-1.32) | 1.04 (0.90-1.22) |
| Comparison of groups | $X^2_{df=3}=22.32$,<br>p<0.001 | $X^2_{df=3}=44.92$,<br>p<0.001 | $X^2_{df=3}=0.26$,<br>p=0.967 | $X^2_{df=3}=2.68$,<br>p=0.443 |
| Age group |  |  |  |  |
| 18-29 | 0.76 (0.64-0.88) | 0.92 (0.85-1.01) | 1.09 (0.95-1.26) | 0.99 (0.91-1.09) |
| 30-49 | 1.12 (1.02-1.21) | 1.06 (1.01-1.12) | 1.06 (0.99-1.15) | 1.05 (0.99-1.12) |
| 50-64 | 1.19 (1.08-1.38) | 1.15 (1.07-1.23) | 1.06 (0.98-1.14) | 1.03 (0.97-1.10) |
| 65+ | 0.97 (0.79-1.20) | 0.96 (0.86-1.07) | 0.97 (1.19-1.12) | 1.02 (0.90-1.15) |
| Comparison of groups | $X^2_{df=3}=34.96$,<br>p<0.001 | $X^2_{df=3}=22.78$,<br>p<0.001 | $X^2_{df=3}=1.94$,<br>p=0.584 | $X^2_{df=3}=1.24$,<br>p=0.743 |
| Education |  |  |  |  |
| <HS graduate | 0.64 (0.54-0.76) | 1.07 (0.99-1.17) | 1.13 (1.00-1.28) | 1.08 (1.00-1.19) |
| HS graduate or GED | 1.25 (1.13-1.36) | 1.14 (1.07-1.21) | 0.99 (0.92-1.07) | 1.02 (0.95-1.08) |
| College, < bachelor's degree | 1.16 (1.07-1.27) | 1.05 (1.00-1.13) | 1.13 (1.04-1.21) | 1.02 (0.95-1.09) |
| Bachelor's degree or more | 0.86 (0.75-0.99) | 0.90 (0.83-0.98) | 1.01 (0.89-1.15) | 1.04 (0.94-1.15) |
| Comparison of groups | $X^2_{df=3}=77.67$,<br>p<0.001 | $X^2_{df=3}=23.37$,<br>p<0.001 | $X^2_{df=3}=8.60$,<br>p=0.035 | $X^2_{df=3}=1.79$,<br>p=0.617 |
| Income |  |  |  |  |
| <\$20K | 1.06 (0.90-1.25) | 1.07 (0.97-1.17) | 1.12 (1.01-1.22) | 1.03 (0.96-1.12) |
| \$20-<\$75K | 1.04 (0.93-1.14) | 1.05 (0.97-1.14) | 1.05 (0.99-1.13) | 1.04 (0.98-1.09) |
| \$75K+ | 1.13 (0.96-1.32) | 1.16 (1.05-1.28) | 0.99 (0.90-1.09) | 1.01 (0.91-1.10) |
| Comparison of groups | $X^2_{df=2}=0.98$,<br>p=0.613 | $X^2_{df=2}=2.61$,<br>p=0.271 | $X^2_{df=2}=2.89$,<br>p=0.235 | $X^2_{df=2}=0.32$,<br>p=0.851 |
| Health insurance |  |  |  |  |
| Private | 1.12 (1.01-1.23) | 1.14 (1.06-1.22) | 0.92 (0.84-1.01) | 1.04 (0.95-1.14) |
| Public | 1.13 (0.95-1.35) | 1.05 (0.95-1.16) | 1.05 (0.91-1.21) | 1.04 (0.94-1.16) |
| None | 0.83 (0.58-1.16) | 1.01 (0.89-1.14) | 1.10 (0.92-1.32) | 1.05 (0.93-1.20) |
| Comparison of groups | $X^2_{df=2}=3.66$,<br>p=0.161 | $X^2_{df=2}=5.31$,<br>p=0.070 | $X^2_{df=2}=4.37$,<br>p=0.113 | $X^2_{df=2}=0.02$,<br>p=0.991 |

a. Coefficients are based on structural equation models adjusting for sex, race/ethnicity, age, education, income, and insurance. In addition, the models adjusted for marital status, employment, country region, and state-level expenditure for tobacco prevention programs compared to CDC recommendations. However, moderating effects were only tested for preselected variables of sex, race/ethnicity, age, education, income, and insurance that were associated with smoking cessation treatment use in past research.

The mediating effects of prescription medications and nicotine replacement therapies appeared to be somewhat larger among past-year smokers with private or public insurance in both 2003 and 2010/11, although these associations did not reach statistical significance.

Analyses of moderated mediation for the association of cigarette taxes with recent smoking cessation in 2010/11 identified few significant effects (Table 4). Counseling and group therapy appeared be a stronger mediator between taxes and recent smoking cessation in middle-aged and older adults; whereas, prescription medications tended to be a stronger mediator in the younger age group. However, this latter association did not reach the predefined p<.01 level of statistical significance (p=0.011). Other noteworthy, but non-significant moderating effects were also found for health insurance: both prescription medications and replacement therapies were stronger mediators of taxes and recent smoking cessation in those with health insurance, whereas the magnitude of mediating effect was larger for counseling and group therapy among the uninsured (Table 4).

**Table 4:** Mediating effect of smoking cessation treatments in the association of cigarette excise taxes and smoking cessation in different socio-demographic groups (moderated mediation), 2010/11.^a^

| 2010/11 survey |  |  |  |
| --- | --- | --- | --- |
| Group | Prescription medications | Replacement Therapies | Counseling |
| Overall | 1.23 (1.15-1.31) | 1.22 (1.15-1.28) | 1.19 (1.05-1.34) |
| Sex |  |  |  |
| Male | 1.20 (1.08-1.32) | 1.21 (1.13-1.30) | 1.17 (0.96-1.43) |
| Female | 1.21 (1.11-1.31) | 1.16 (1.07-1.26) | 1.28 (1.08-1.52) |
| Comparison of groups | $X^2_{df=2}=0.01, p=0.923$ | $X^2_{df=2}=0.76, p=0.383$ | $X^2_{df=2}=0.82, p=0.364$ |
| Race/ethnicity |  |  |  |
| Non-Hispanic white | 1.20 (1.10-1.30) | 1.15 (1.07-1.22) | 1.25 (1.06-1.48) |
| Non-Hispanic black | 1.10 (0.88-1.38) | 1.36 (1.20-1.55) | 1.12 (0.84-1.48) |
| Hispanic | 1.58 (1.16-2.14) | 1.35 (1.10-1.63) | 1.40 (0.83-2.39) |
| Other | 1.14 (0.76-1.70) | 1.21 (0.96-1.54) | 1.26 (0.69-2.27) |
| Comparison of groups | $X^2_{df=3}=4.12, p=0.249$ | $X^2_{df=3}=7.02, p=0.071$ | $X^2_{df=3}=1.06, p=0.788$ |
| Age group |  |  |  |
| 18-29 | 1.54 (1.27-1.86) | 1.14 (0.99-1.30) | 1.31 (1.01-1.68) |
| 30-49 | 1.21 (1.07-1.36) | 1.23 (1.13-1.34) | 0.94 (0.76-1.16) |
| 50-64 | 1.13 (1.02-1.23) | 1.16 (1.06-1.27) | 1.48 (1.22-1.79) |
| 65+ | 1.06 (0.90-1.25) | 1.20 (1.03-1.39) | 1.62 (1.13-2.29) |
| Comparison of groups | $X^2_{df=3}=11.20, p=0.011$ | $X^2_{df=3}=1.52, p=0.677$ | $X^2_{df=3}=14.07, p=0.003$ |
| Education |  |  |  |
| <HS gradate | 1.35 (1.13-1.63) | 1.19 (1.04-1.34) | 1.25 (0.96-1.62) |
| HS graduate or GED | 1.10 (0.99-1.22) | 1.16 (1.06-1.28) | 1.10 (0.86-1.43) |
| College, < bachelor's degree | 1.23 (1.10-1.38) | 1.25 (1.14-1.36) | 1.36 (1.12-1.67) |
| Bachelor's degree or more | 1.25 (1.04-1.49) | 1.14 (1.01-1.28) | 1.19 (0.91-1.52) |
| Comparison of groups | $X^2_{df=3}=5.32, p=0.150$ | $X^2_{df=3}=1.77, p=0.621$ | $X^2_{df=3}=2.16, p=0.541$ |
| Income |  |  |  |
| <\$20K | 1.23 (1.08-1.40) | 1.27 (1.16-1.40) | 1.21 (0.93-1.57) |
| \$20-<\$75K | 1.25 (1.14-1.36) | 1.14 (1.05-1.23) | 1.25 (1.05-1.48) |
| \$75K+ | 1.05 (0.90-1.21) | 1.21 (1.08-1.36) | 1.27 (1.01-1.62) |
| Comparison of groups | $X^2_{df=2}=5.69, p=0.058$ | $X^2_{df=2}=3.87, p=0.144$ | $X^2_{df=2}=0.13, p=0.939$ |
| Health insurance |  |  |  |
| Private | 1.35 (1.19-1.55) | 1.30 (1.16-1.46) | 1.21 (0.91-1.58) |
| Public | 1.42 (1.15-1.77) | 1.49 (1.31-1.72) | 0.95 (0.68-1.34) |
| None | 1.17 (0.88-1.57) | 1.10 (0.91-1.34) | 1.54 (1.04-2.29) |
| Comparison of groups | $X^2_{df=2}=1.20, p=0.548$ | $X^2_{df=2}=7.93, p=0.019$ | $X^2_{df=2}=3.99, p=0.136$ |

a. Coefficients are based on structural equation models adjusting for sex, race/ethnicity, age, education, income, and insurance. In addition, the models adjusted for marital status, employment, country region, and state-level expenditure for tobacco prevention programs compared to CDC recommendations. However, moderating effects were only tested for preselected variables of sex, race/ethnicity, age, education, income, and insurance that were associated with smoking cessation treatment use in past research.

## Discussion

We examined whether the mediating effect of smoking cessation treatment use between tobacco policies and smoking cessation varied across different socio-demographic groups in the 2003 and 2010/11 CPS-TUS. For clean indoor air laws, the mediating effect of prescription medications and nicotine replacement therapies varied significantly between racial/ethnic, age, and education groups in 2003. However, none of these moderation effects remained significant in 2010/11. For cigarette excise taxes, the mediating effect of counseling was significantly moderated by age groups in 2010/11 and the same moderator showed a trend-level significance for prescription medications. No other candidate moderator reached statistical significance.

We found significant differences in smoking cessation treatment use by sociodemographic background. Consistent with existing research based on PATH data (19), African American and Hispanic smokers were less likely to use evidence-based smoking cessation treatments in both 2003 and 2010/11 and the same was the case for young adult smokers. The low adoption of evidence-based smoking cessation strategies may explain why African American smokers try to quit more frequently than Whites, but are less likely to achieve abstinence (29). In contrast to other studies, our analyses of CPS-TUS data showed higher treatment use among women and those with higher education, which has not been found in more recent PATH data (19). Findings of the current study confirm that additional efforts to reach young people and racial/ethnic minorities with evidence based smoking cessation strategies are needed.

Moreover, smokers without health insurance had a lower likelihood to use any type of smoking cessation treatment, including treatments like quitlines and internet resources, which are typically free of charge. This could suggest that in addition to treatment costs, there may be other barriers to treatment access, for example related to knowledge, attitudes, and social norms among uninsured smokers. Moving forward, it will be important to use later waves of the CPS-TUS to investigate the impact of the 2010 Patient Protection and Affordable Care Act (ACA), which required many public and private insurers to cover all FDA approved cessation medications and counselling without insurance barriers (1).

Our findings also demonstrated that sociodemographic moderators of the association between tobacco policies and smoking cessation decreased from 2003 to 2010/11, and this was observed for both cigarette taxes and clean indoor air laws. These results extend earlier findings suggesting that the impact of tobacco control policies on population level smoking cessation is, in part, effected through stimulating the use of evidence-based smoking cessation treatments. In recent years, this pathway between policies and smoking cessation does not seem to differ in magnitude across various sociodemographic groups. Existing research has demonstrated a pro-equity impact of tobacco taxes on smoking disparities by socioeconomic status (30,31). The evidence for clean indoor air laws in reducing smoking inequities is less clear (31), but some studies suggest that nationwide and comprehensive clean indoor air laws may have a more positive impact on equity compared to regional and voluntary policies (30). Together with this exsiting literature, our findings suggest that tobacco control policies can contribute to a reduction in tobacco-related health disparities. In order to further reduce these disparities, an emphasis on policies that remove barriers to smoking cessation treatment access among vulnerable populations may be needed. For example, the elimination of co-payments for smoking cessation medication has been associated with an increase in medication use, particularly among low-SES smokers (32).

## Limitations

Our analysis has limitations. We assessed the association of taxes and clean indoor air laws with recent smoking cessation and the results may be different for long-term cessation. Tobacco control policies may have a delayed effect on smoking cessation (33), yet our data were limited to past year smoking behaviour and we could not track the association of policies with smoking cessation over time. In addition to smoking cessation treatment use, as investigated in the current study, other behaviours or attitudes may also act as mediators between tobacco policies and smoking cessation. These other factors were subsumed in the “direct effects” in our mediation analysis and further studies are needed to explore their potential contribution. Due to the strong correlation between clean indoor air laws and cigarette taxes, we were not able to assess their effects in models including both variables. Finally, data analyzed were from 2003 and 2010/11 and it is unclear if findings hold true today. Updated analyses including the latest wave of CPS-TUS data are warranted moving forward.

## Conclusions

Sociodemographic differences in the effect of clean indoor air laws and excise taxes on smoking cessation treatment use have decreased from 2003 to 2010/11. Although we found some evidence of moderation by sociodemographic characteristics, in most cases, tobacco control policies appear to impact smoking cessation by stimulating smoking cessation treatment use similarly across groups. Taken together, our findings support the expansion of tobacco control policies, including cigarette taxes and clean indoor air laws, and continued investment in evidence-based smoking cessation treatments, to further reduce smoking in the US population. Such tobacco control efforts appear to be efficacious in reducing smoking among vulnerable groups, including young people, racial/ethnic minorities, and those with low education or income. However, additional efforts are needed to promote smoking cessation in these high priority groups for a pro-equity impact of tobacco control policies and close the gap in smoking cessation rates across population subgroups.

## Data Availability

CPS-TUS data are available from https://cancercontrol.cancer.gov/brp/tcrb/tus-cps/questionnaires-data.

